# Knowledge of healthcare workers in the domain of oncogenetics

**DOI:** 10.1101/2024.11.27.24318049

**Authors:** Jad Jabbour, Léa Habibian, Christine Anne-Marie Martin, Ernest Diab, Hampig Raphaël Kourié

**Affiliations:** Department of Hematology and Oncology, Hôtel Dieu de France University Hospital, Saint Joseph University of Beirut, Beirut, Lebanon

**Keywords:** Oncogenetics, Cancer, Genetic testing, Genetic counseling, Healthcare workers, Breast cancer, Knowledge assessment, medical education, Awareness

## Abstract

**Background:** Cancer is the second leading cause of mortality globally. 5-10% of cancer cases involve a genetic factor. Oncogenetics studies the role of genetic mutations in the development of cancer and plays a crucial role in understanding cancer pathogenesis and developing targeted therapies. Despite the critical nature of this field, the knowledge level among healthcare workers regarding oncogenetics, particularly in breast cancer, remains underexplored.

**Objective:** This study aimed to assess the knowledge of healthcare workers in oncogenetics, with a specific focus on breast cancer, to identify gaps and potential areas for educational intervention.

**Methods:** A cross-sectional, web-based survey was conducted at a University Hospital. A questionnaire consisting of general and breast cancer-specific oncogenetics questions was distributed to nurses, medical interns, residents, and doctors. Responses were scored and analyzed to potentially identify statistically significant differences based on professional roles and experience.

**Results:** 184 answers were recorded from which 89.67% confirmed familiarity with the term oncogenetics. Medical interns, residents, and doctors demonstrated significantly higher knowledge compared to nurses (p<0.001). The average scores indicated better general oncogenetics knowledge (mean=4.83/6) than breast cancer-specific knowledge (mean=3.77/6). Familiarity with oncogenetics correlated with higher scores across both sections (p<0.001). Notably, male participants outperformed females in breast cancer oncogenetics (p=0.02).

**Conclusion:** Healthcare workers displayed a satisfactory general knowledge of oncogenetics but showed significant gaps in breast cancer-specific knowledge. These findings highlight the need for targeted educational programs to enhance oncogenetics competency among healthcare workers, ensuring improved patient care. Future studies should evaluate the impact of such educational interventions on healthcare workers’ knowledge and practice.

## Introduction

Cancer is the second leading cause of mortality worldwide accounting for around 10 million deaths in 2020 (1). Factors such as genetics, age, sex, behaviors like tobacco use and alcohol consumption, and history of infections can increase the risk of developing cancer (1-2). In 2018, 13% of cancers diagnosed globally were attributed to carcinogenic infections, such as *Helicobacter pylori*, Hepatitis B and C viruses, and Epstein-Barr virus (1). Although 90 to 95% of cancer cases are linked to environmental factors, 5-10% of all cancer cases can be associated with genetic defects (2). Cancer results from the alteration of important regulatory factors that control cell proliferation, differentiation, and apoptosis, such as oncogenes and tumor suppressor genes (3). Mutations can take two different paths: they can either act like a pedal in a car, accelerating cell division, or, conversely, act as a brake, also resulting in neoplasm (4).

More than 40 years ago, the first proto-oncogene discovered was the SRC gene, derived from the Rous sarcoma virus (RSV), which helped researchers understand the mechanisms of oncogenes and their therapeutic implications (5). A proto-oncogene is a cell regulatory gene that mainly encodes proteins responsible of signal transduction pathways, therefore controlling the normal cell proliferation. However, an oncogene is the result of an abnormal expression or a mutation of a corresponding proto-oncogene, which induces abnormal cell proliferation and tumor development (6-7). Oncogenes can be activated in three ways: point mutation, gene amplification, and chromosomal rearrangement (8).

With the increasing importance of genetics in the pathogenesis of cancer, the number of genetic counselors has risen globally. They provide genetic counseling sessions, serving as a vital resource of information regarding genetic disorders for healthcare professionals, patients, and the public (9). These sessions aim to support the public in understanding genetic conditions, discussing associated risks, exploring further testing, and managing these conditions medically and psychologically (9-10).

The need for genetic testing and counseling can be identified by primary care providers, oncology specialists, or cancer genetics professionals. However, some genetic testing options are also available to anxious consumers wishing to take control of their genetic health (10). Moreover, collected genetic information play a major role in oncology care, surgery and targeted chemotherapy (10-11). Gathering family history information is the initial step in determining who would benefit from genetic testing, tailored screening, and risk-reducing interventions. Consequently, professional societies have published guidelines to assist clinicians in identifying individuals who should be offered genetic risk assessment and testing (12).

In today’s practice, general practitioners are confronted to challenging situations in regards of genetic information, patients’ requests for genetic tests, their diagnostic value and therapeutic consequences. However, studies have shown that general practitioners’ knowledge and competencies in oncogenetics remain unclear among non-geneticist healthcare workers. For them to contribute effectively in the field of oncogenetics, their knowledge needs to be upgraded (13-14).

The discovery of some mutations of oncogenes has led to changes in the treatment regimens of certain types of cancer through targeted therapies and their impact on the prognosis of cancer previously defined as incurable cancers (11). For instance, the BCR/ABL1 oncogene, involved in the development of chronic myeloid leukemia (CML), is inhibited by the tyrosine kinase inhibitor Imatinib. Following its discovery, CML, once considered a fatal cancer, is now regarded as a chronic disease (15-16). Around 30% of all human tumors are linked to a mutation in the *Ras* genes (17). Moreover, HER2, a membrane tyrosine kinase and an oncogene, is expressed in approximately 15 to 20% of breast tumors and can be targeted by Trastuzumab, also known as Herceptin (18). Thus, the contribution of oncogenetics in the field of oncology is undeniably increasing.

## Aim of the study

Acknowledging the importance of this topic in oncology, we were curious to be acquainted with the knowledge of the healthcare providers in the domain of oncogenetics in general and regarding breast cancer specifically. The motive behind choosing breast cancer is its prevalence as the most common type of cancer in the world and Lebanon specifically (20-21-22). Consequently, we conducted a study to assess the level of awareness and knowledge of nurses, medical interns, residents, and doctors. Indeed, understanding the cellular and genetic mechanisms in certain types of cancers is the key for prevention, diagnosis, and targeted treatments for the patient. Thus, our goal is to be able to evaluate the knowledge of health professionals in the domain of oncogenetics with the possibility of developing a new management model. The latter includes various health professionals aiming to reduce the demand for specialists in genetics while respecting professional requirements, ethical and legal practice of genetics. This study’s results will also evaluate the importance of the implementation of educational programs and workshops to boost the knowledge and the practice of oncogenetics to boost the practice of oncogenetics and make it a routine step in the personalized care of each patient with the new life-saving treatments.

## Material and methods

This is a web-based, cross-sectional descriptive study. A questionnaire was made from scratch and distributed to healthcare professionals in a university hospital (Appendix 1). The questionnaire targeted nurses, medical interns, residents, and doctors (MD) practicing along patients in the hospital.

The questionnaire was divided into three sections. The first aimed to gather background information about the respondents, including their role in the hospital, age, years of experience, and familiarity with the term ‘oncogenetics.’ The second section comprised six questions designed to assess the respondents’ general knowledge of oncogenetics. Finally, the last part also containing 6 questions, was more targeted towards breast cancer oncogenetics as it is the most encountered types of cancer (20-21-22). Each question in the last 2 sections had 4 choices of answers but only one of them was the right answer. The questionnaire was sent via Google Forms and answers were collected on the same platform. The results were analyzed using R software. For ease of analysis, each correct answer was assigned 1 point, while incorrect answers were given 0 points. The total points from the first and second sections, as well as the overall questionnaire, were used for data analysis. The maximum possible score for each section was 6 points, with a total of 12 points for the entire questionnaire. Data that showed a sample size less than 30, were evaluated under the Shapiro-Wilk test to test their normality. Student’s T-test and one-way ANOVA test were used to compare means when data were considered normally distributed or had a sample size bigger than 30. The Mann-Whitney U test and the Kruskal-Wallis test were used when data did not meet the previous normality criteria. The statistical tools included parameters with a confidence level of 95%. Analysis showing a p-value <0.05 according to Cronbach’s alpha was deduced to be statistically significant.

### Ethical considerations

This study was conducted in compliance with the ethical principles outlined in the Declaration of Helsinki and approved by the institutional ethics review board of Université Saint-Joseph de Beyrouth. Participants were provided with detailed information about the study’s purpose, procedures, and their rights prior to participation. By voluntarily answering the online questionnaire, participants indicated their informed consent to participate in the study. Participation was entirely voluntary, and participants were informed of their right to withdraw at any time without any consequences. The study did not include minors; all participants were healthcare professionals aged 18 or older. As the study involved a web-based survey, no personally identifiable information was collected to ensure participant anonymity and confidentiality.

## Results

A total of 184 participants answered the questionnaire. Table 1 summarizes characteristics of the participants.

**Table 1:** Characteristics of the participants.

| **Characteristics** | **Count (%)**<br>*N=184* |
| --- | --- |
| **Sex** |  |
| Male | 86 (46.74%) |
| Female | 98 (53.26%) |
| **Age** |  |
| 18-30 | 146 (79.35%) |
| 30-40 | 14 (7.61%) |
| 40-50 | 10 (5.43%) |
| >50 | 14 (7.61%) |
| **Function within the hospital** |  |
| Nurse | 36 (19.57%) |
| Medical Intern | 77 (41.85%) |
| Medical Resident | 49 (26.63%) |
| MD specialist | 22 (11.96%) |
| **Years of experience** |  |
| <5 years | 123 (66.85%) |
| 5-10 years | 32 (17.39%) |
| >10 years | 29 (15.76%) |
| **Familiarity with the term oncogenetics** |  |
| Yes | 165 (89.67%) |
| No | 19 (10.33%) |

Female participants responded slightly more than male participants, resulting in a sex ratio of 0.88 M/F. The majority of respondents were in the 18-30 age group, accounting for 79.35% of participants. Among the 184 participants, medical interns constituted the largest group, with 77 (41.85%) participants, followed by medical residents with 49 (26.63%), nurses with 36 (19.57%), and MD specialists with 22 (11.96%). Medical residents and specialists had backgrounds in various fields, including internal medicine, surgery, orthopedics, pediatrics, and hematology-oncology. Regarding years of experience, the majority of respondents, totaling 123 (66.85%), had less than 5 years of experience. In comparison, 32 (17.39%) participants reported having between 5 and 10 years of experience, while 29 (15.76%) had more than 10 years of experience. Additionally, 165 (89.67%) participants claimed to be familiar with the term oncogenetics. Among the MD specialists, 13 (59.09%) indicated that they had referred at least one of their patients to an oncogenetics specialist, whereas 9 (40.91%) had requested oncogenetic testing themselves (Figure 1).

**Figure 1:**
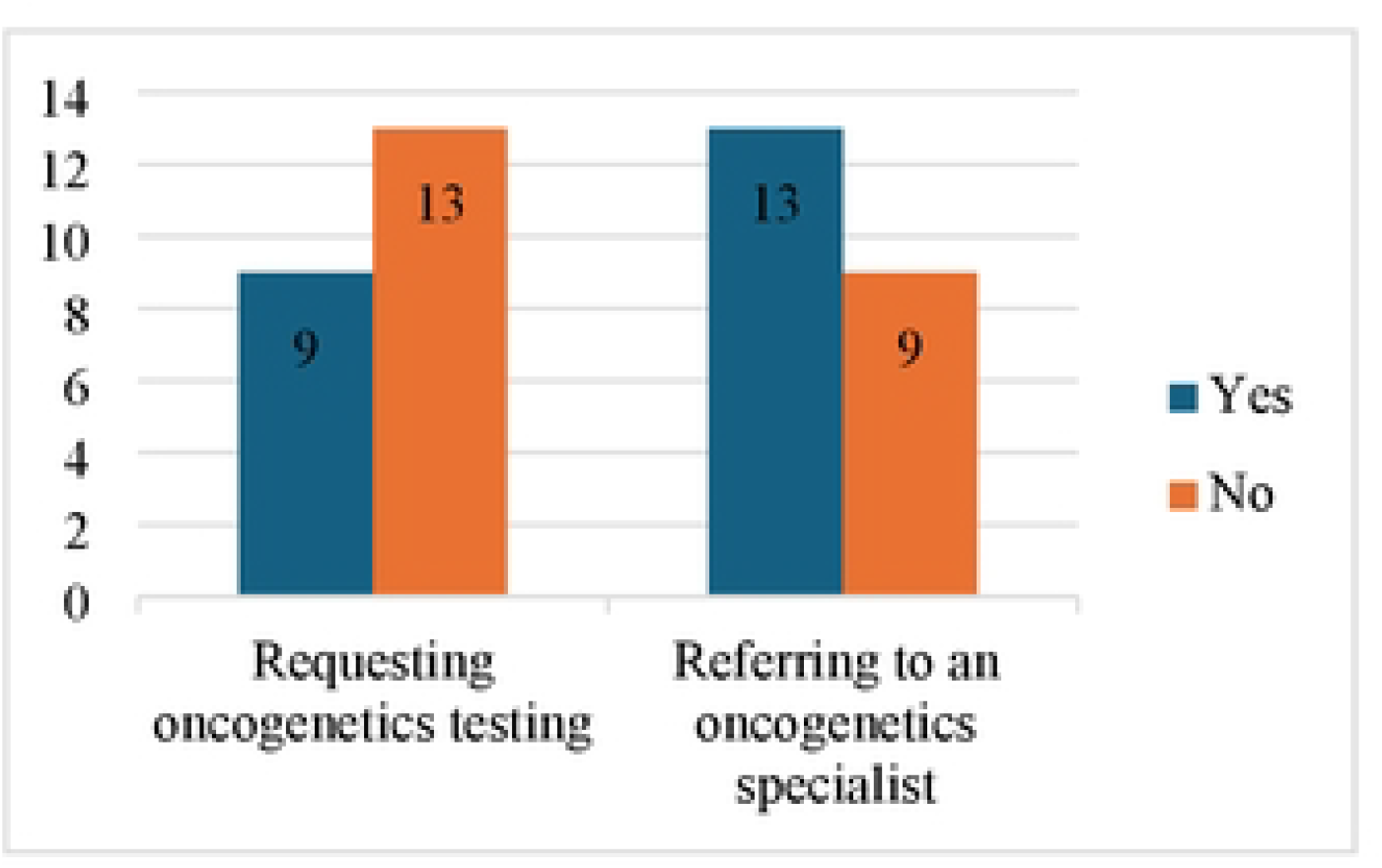
Practice of the oncogenetics domain among MD specialists

After attributing a score of 1 point to each correct answer and 0 for wrong answers, the questions and number of correct answers are presented in table 2 for the first section and table 3 for the second section.

**Table 2:**
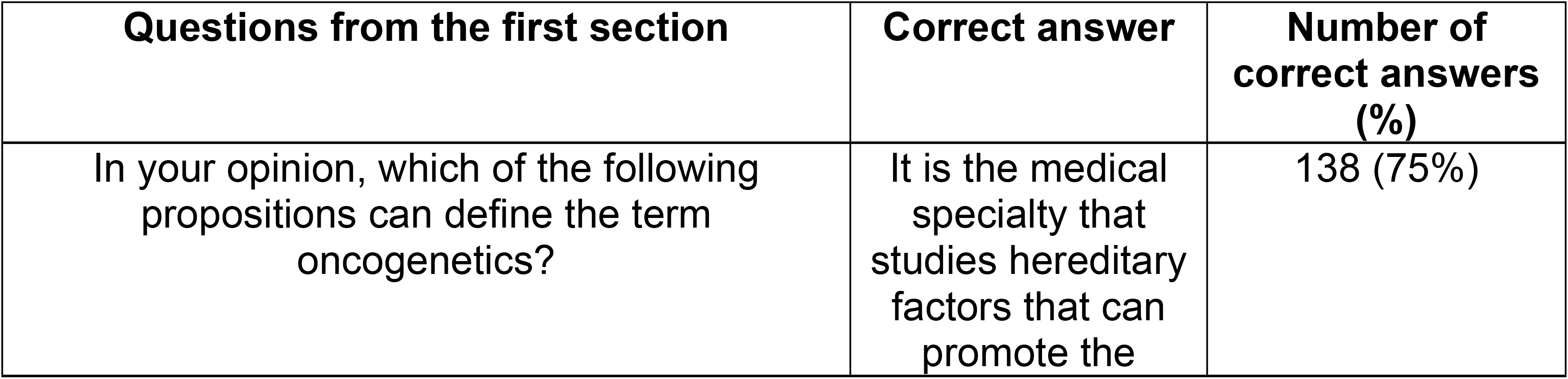
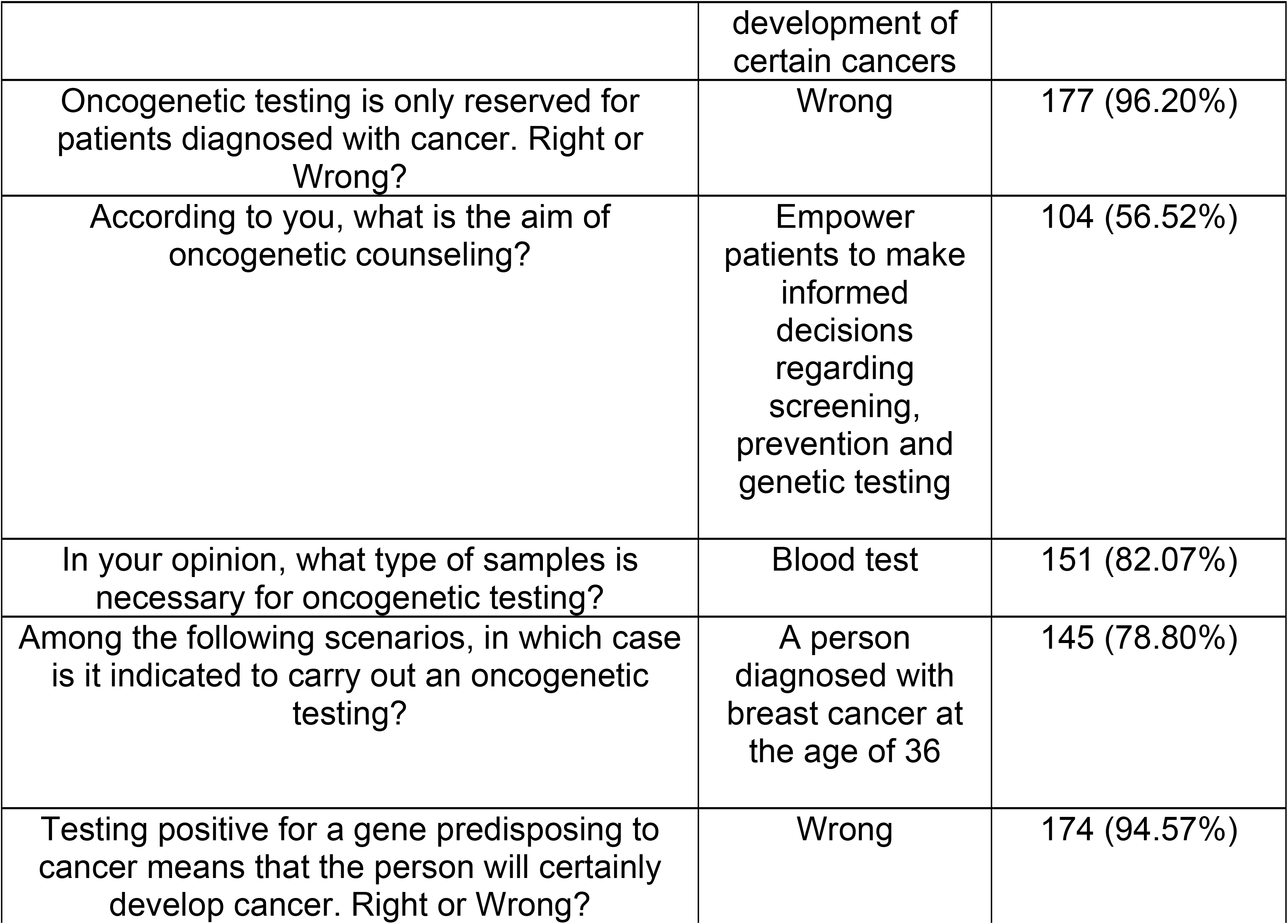
Count of right and wrong answers to each question of the first section of the questionnaire.

**Table 3:** Count of right and wrong answers to each question of the second section of the questionnaire.

| **Questions of the second section** | **Correct answer** | **Number of correct answers (%)** |
| --- | --- | --- |
| Which of these genes is the most predisposing to developing breast cancer? | BRCA1-2 | 175 (95.11%) |
| Which gene playing a role in the oncogenetics of breast cancer is most associated with a risk of ovarian cancer? | BRCA1 | 56 (30.43%) |
| Having BRCA1 or BRCA2 gene means: | The patient is more likely to develop breast cancer at a younger age | 176 (95.65%) |
| Concerning BRCA1 and BRCA2 genes, choose the right answer: | These are tumor suppressor genes | 91 (49.46%) |
| Regarding the genetic testing of BRCA1-2 genes, choose the right answer: | Negativity of the test ensures non-transmission of the gene to offspring | 45 (24.46%) |
| Regarding cancers with a mutated BRCA gene, choose the right answer: | BRCA mutation is associated with the development of breast cancer at a younger age | 150 (81.52%) |

Based on the answers of the first questionnaire, the lowest score was obtained in the questioning discussing the aim of oncogenetic counseling (56.52%). However, a relatively high proportion of participants correctly answered the other questions of this section.

The second section of the questionnaire focused on the oncogenetics of breast cancer. Most questions pertained to facts about breast cancer and oncogenetics. Participants demonstrated a lack of knowledge regarding the nature of the BRCA1 and BRCA2 genes, with only 49.46% answering correctly. Additionally, only 30.43% identified which gene is most implicated in the genomics of breast cancer, and just 24.46% understood how to interpret an oncogenetic test result. Averages of scores were calculated regarding different variables and are represented in table 4.

**Table 4:** Average scores of participants in the first, second sections and total questionnaire according to their function in the hospital, sex and familiarity with the term oncogenetics.

| **Variable** | **First section**<br>**(mean±SD)**<br>**Score /6** | **Second section**<br>**(mean±SD)**<br>**Score /6** | **Overall score**<br>**(mean±SD)**<br>**Score /12** |
| --- | --- | --- | --- |
| **Function within**<br>**the hospital** |  |  |  |
| Nurse | 3.97 ± 1.34 | 3.03 ± 1.13 | 7.00 ± 1.91 |
| Medical Intern | 5.06 ± 0.91 | 3.96 ± 0.87 | 9.03 ± 1.35 |
| Medical Resident | 5.02 ± 0.99 | 3.88 ± 0.88 | 8.90 ± 1.42 |
| MD specialist | 4.95 ± 1.00 | 4.05 ± 1.21 | 9.00 ± 1.69 |
| **Sex** |  |  |  |
| Male | 4.85 ± 1.05 | 3.95 ± 0.92 | 8.80 ± 1.54 |
| Female | 4.81 ± 1.17 | 3.60 ± 1.10 | 8.41 ± 1.84 |
| **Familiarity with**<br>**the term**<br>**oncogenetics** |  |  |  |
| Yes | 4.93 ± 1.06 | 3.89 ± 0.93 | 8.82 ± 1.55 |
| No | 3.95 ± 1.22 | 2.68 ± 1.25 | 6.63 ± 1.83 |
| **Total** | 4.83 ± 1.11 | 3.77 ± 1.03 | 8.59 ± 1.72 |

The 184 participants averaged 4.83 ± 1.11 points on the first section, 3.77 ± 1.03 on the second section, and 8.59 ± 1.72 on the overall questionnaire. The comparison of results from the first and second sections using the Student’s T-test revealed a statistically significant difference (p < 0.001), indicating better performance in general oncogenetics knowledge compared to breast cancer oncogenetics.

Based on the Kruskal-Wallis test, the average scores obtained in the first section of the questionnaire showed a statistically significant difference when comparing nurses’ scores with those of medical interns (p < 0.001), medical residents (p < 0.001), and MD specialists (p < 0.001). However, there was no statistically significant difference in scores between medical interns, residents, and specialists in the first section.

Regarding the second section of the questionnaire and the overall questionnaire scores, the one-way ANOVA analysis for each, showed the same result with a statistically significant difference between nurses on one side, and medical interns, medical residents, and MD specialists on the other (p<0.001). When comparing scores obtained from female and male participants, the Student’s T-test did not show a statistically significant difference in the first section (p = 0.79) or in the overall score (p = 0.12). However, the average scores in the second section, which focused on the oncogenetics of breast cancer, showed a statistically significant difference (p = 0.02) in favor of the males. Nineteen participants (10.33%) claimed not to be familiar with the term oncogenetics. Among them, 16 (84.21%) were nurses, 2 were residents (one in family medicine and one in emergency medicine), and 1 was a specialist in pediatrics.

The comparison of average scores obtained from familiar participants and non-familiar participants with oncogenetics were conducted using the Mann-Whitney U test (first section and the overall score) and the Student’s T-test (second section). The comparison was statistically significant when comparing the scores of the first section (p<0.001), the second section (p<0.001) and the overall score (p<0.001).

## Discussion

The results obtained showed decent knowledge of oncogenetics among healthcare professionals in the tertiary hospital. The latter, being a university teaching hospital, explains the relative predominance of participants enrolled in the medical education program, therefore behind the highest proportion of participants being between 18 and 30 years old. People who go through medical school (medical interns, residents and specialists) show a higher level of knowledge compared to nurses. This result could be related to the exposure to oncogenetics whether during the medical studies or during practice of medicine. In fact, in a study conducted by Prolla CM et al. in 2015, nurses presented some lack of knowledge in the oncogenetics of breast cancer and concluded the need of actions to be planned in order to reinforce controlling breast cancer (23). Hébert J et al. have demonstrated that even nurses in an oncology setting tend to lack some knowledge about oncogenetics which could affect their practice especially when assessing the patients (24). Whether it’s nurses of medical graduates, the need of educational programs is important because healthcare workers play an important role in the prevention and the spread of information among the population in regard to breast cancer (23-24). This problem has existed since 2001 and is still ongoing nowadays (25). In a recent study in Saudi Arabia in 2023, almost half the population showed poor knowledge of cancer genetics and the importance of genetic testing (26). The latter could be the case of the Lebanese population, therefore a good knowledge of healthcare workers is required to increase the general population’s knowledge of cancer genetics and testing. Educational and training interventions play an important role in enhancing the knowledge of healthcare workers in the domain of oncogenetics. This statement was deduced already by Houwink EJ et al. where an educational intervention seemed beneficial for general practitioners in the domain of oncogenetics (14). Even though participants in this study showed an overall satisfying level of knowledge in the domain of oncogenetics, educational interventions and trainings are still needed to improve their knowledge in this domain for a better practice of medicine. When comparing sections of the study, difference in performance between both seemed evident. Healthcare workers demonstrated lower levels of knowledge in breast cancer oncogenetics even if it was the most common cancer in Lebanon and worldwide (20-21-22). Nurses appear to be less familiar with the term ‘oncogenetics.’ Of the 19 participants unfamiliar with the term, 16 were nurses. A comparison of the scores revealed significantly higher results among those familiar with oncogenetics, suggesting that score performance could reflect familiarity with the term. Male participants, on the other hand, demonstrated better knowledge in breast cancer oncogenetics, and this difference in performance was found to be statistically significant. However, this finding may be influenced by the gender distribution within the nurse group, where 29 of the nurses were female and only 7 were male. Additionally, 16 of the 19 participants unfamiliar with the term were female. Given the lower knowledge level demonstrated by nurses and the number of females unfamiliar with oncogenetics, the assumption about gender differences in knowledge should be interpreted with caution and requires further confirmation. To our knowledge, this type of study is the first conducted in Lebanon assessing the healthcare workers’ knowledge in the domain of oncogenetics and breast cancer genetics specifically. The questionnaire that was distributed is a small simple questionnaire, reducing the possible biases that could be generated by long questionnaire due to loss of concentration. The questionnaire contains questions that could be encountered daily by any healthcare worker, therefore it is testing the spread of correct information among the general population. Our sample is a large sample from a single tertiary hospital. It can represent any teaching facility where young practitioners are predominant like in our sample and healthcare workers in general in Lebanon. The study being conducted in a single center in Lebanon cannot be representative of the Lebanese population in general and is only representing an institution’s knowledge. The latter could affect the transferability of the results obtained in this study. The questionnaire that was distributed among the healthcare workers was established by the researchers to test the knowledge of healthcare workers in the domain of oncogenetics. However, for better assessment of knowledge, we need a universal version of the questionnaire.

## Conclusion

Oncogenetics has been reshaping the field of cancer in recent years, offering tools that are vital for the diagnosis and treatment of the disease. Assessing the knowledge of healthcare workers in oncogenetics is important for improving cancer prevention among the entire population. Overall, healthcare workers demonstrate a good understanding of oncogenetics; however, nurses tend to have a relatively lower level of knowledge compared to medical interns, residents, and specialists. Despite breast cancer being the most commonly diagnosed cancer in Lebanon and worldwide, there seems to be a knowledge gap specifically related to its oncogenetics. Healthcare workers are often the primary source of information for the general population regarding cancer and genetics. Therefore, educational interventions aimed at both the general public and healthcare workers are warranted. Future studies should evaluate the effectiveness of these interventions in enhancing the knowledge of healthcare workers in the field of oncogenetics.

## Data Availability

All relevant data are within the manuscript and its Supporting Information files.

## Notes

### Competing Interest Statement

The authors have declared no competing interest.

### Funding Statement

The author(s) received no specific funding for this work.

### Author Declarations

This study was conducted in compliance with the ethical principles outlined in the Declaration of Helsinki and approved by the institutional ethics review board of Université Saint-Joseph de Beyrouth.

